# DermAssist: A Hybrid Vision Transformer System for Skin Lesion Diagnosis with Automated Alerting and Dual-Sided Portals

**DOI:** 10.1101/2025.07.15.25331570

**Authors:** Safwan Nasir

## Abstract

Skin cancer, one of the most prevalent forms of cancer globally, demands early and accurate diagnosis to improve patient outcomes. In this paper, we present **DermAssist**, a hybrid deep learning-based dermatology assistant system that integrates Convolutional Neural Networks (CNNs) and Vision Transformers (ViTs) for automated multi-class skin lesion classification. Our model was trained and evaluated on widely-used datasets including DermNet, ISIC, and HAM10000, employing a robust preprocessing pipeline to enhance lesion visibility and diversity. DermAssist combines segmentation (U-Net), feature extraction (ResNet), and classification (BEiT Transformer), and introduces a dual-portal architecture for clinicians and patients using Streamlit and Flask interfaces. We incorporate Grad-CAM and SHAP for interpretability, Twilio-based SMS alerting for high-risk cases (*>*90% confidence), and secure AWS S3 storage to ensure HIPAA compliance. Experimental results demonstrate an accuracy of 90.2% with strong ROC-AUC and precision scores. DermAssist is positioned as a deployable, intelligent diagnostic aid capable of enhancing dermatological workflows in real-time environments.

## I. Introduction

Skin cancer continues to be one of the most common cancers worldwide, with millions of new cases diagnosed annually. The rising incidence of melanoma, basal cell carcinoma (BCC), and squamous cell carcinoma (SCC) has placed a growing burden on healthcare systems, especially in regions with limited access to dermatologists. Early detection is critical for increasing survival rates, yet traditional diagnostic workflows are manual, time-intensive, and subject to variability in clinical expertise.

With the advent of artificial intelligence (AI), computer-aided diagnosis (CAD) systems are being increasingly adopted for dermatological screening. These systems aim to assist clinicians by providing consistent and accurate lesion analysis using deep learning algorithms. However, challenges such as false positives, lack of interpretability, inadequate deployment pipelines, and regulatory concerns still hinder their real-world adoption.

To address these issues, we propose **DermAssist**, an end-to-end intelligent assistant that leverages the power of hybrid deep learning architectures and automated alerting to facilitate early skin cancer diagnosis.

### A. Background and Motivation

Dermatologists often rely on the ABCDE rule (Asymmetry, Border, Color, Diameter, and Evolving) for evaluating lesion malignancy. While dermoscopy improves accuracy, access to specialized equipment and trained professionals remains a barrier in many healthcare systems. Automated diagnostic tools, especially those powered by deep learning, can democratize skin cancer screening by enabling real-time, scalable, and accurate lesion evaluation.

Recent advances in neural networks have yielded models like VGGFace, MobileNet, and BEiT that generalize well across diverse domains. However, dermatological imaging presents challenges such as low contrast, lesion occlusion, inter-class similarities, and the need for explainability. Integrating CNN-based encoders, ViT classifiers, and attention-based interpretability methods like Grad-CAM and SHAP can significantly enhance diagnostic reliability and trust.

### B. Problem Statement

Despite promising results in skin lesion classification using CNNs, most existing systems suffer from:

- Limited spatial awareness due to local convolutional filters.
- Lack of real-time diagnosis and automated clinical alerting.
- Weak interpretability and clinical transparency in decision-making.
- Vulnerabilities to occlusion, lighting variations, and dataset biases.

Moreover, privacy and regulatory requirements, including HIPAA compliance, are rarely addressed in academic prototypes.

### C. Objectives and Scope

This project aims to build a hybrid AI system, DermAssist, capable of:

- Diagnosing seven major skin lesion types from dermoscopic images.
- Combining U-Net for lesion segmentation, ResNet for feature extraction, and BEiT for classification.
- Providing explainable predictions via Grad-CAM and SHAP.
- Sending automated alerts via Twilio when high malignancy confidence is detected.
- Delivering a dual-interface web application for patients and doctors via Streamlit and Flask.
- Hosting encrypted diagnostic data on AWS S3, ensuring HIPAA compliance.

The system is intended for use in clinical and remote telemedicine settings, with modularity for future multimodal (e.g., image + text) expansions.

### D. Key Contributions

This paper makes the following contributions:

1. **Hybrid Model Architecture:** We present a multi-stage system using U-Net, ResNet, and BEiT for end-to-end lesion segmentation, feature learning, and classification.
2. **Real-Time Clinical Alerts:** DermAssist includes a high-risk alert module integrated with Twilio for immediate SMS notifications to clinicians.
3. **Dual-Sided Portal Deployment:** We design Streamlit (patient) and Flask (doctor) interfaces for seamless diagnostic access, hosted securely with AWS S3 encryption.
4. **Interpretability Layer:** Grad-CAM and SHAP visualizations are incorporated to highlight decision-making regions and support clinical trust.
5. **Robust Evaluation:** We report comprehensive metrics (accuracy, ROC, confusion matrix) using DermNet, ISIC, and HAM10000 datasets, showing 90.2% accuracy.

### E. Clinical Relevance and Use Case

Skin cancer diagnosis often begins in primary care, where access to specialists is limited. A system like DermAssist allows early-stage triaging by non-specialist physicians and self-screening by patients. Its ability to flag high-confidence malignant cases (*>* 90%) for immediate alerts and provide web-based diagnostics portals ensures timely medical intervention and broader outreach.

### F. Dataset Diversity and Bias Mitigation

To ensure generalizability, we curated and trained on diverse datasets—HAM10000, DermNet, and ISIC—that include varied skin tones, lesion types, and imaging conditions. Augmentation techniques like random cropping, histogram equalization, and color jittering were employed to simulate real-world imaging variability and reduce dataset-induced bias.

### G. Privacy and Regulatory Compliance

DermAssist enforces Health Insurance Portability and Accountability Act (HIPAA) standards via:

- Encrypted image storage using Amazon S3 with access control lists (ACLs).
- Deletion of raw images post-feature extraction.
- Secure login mechanisms for clinicians and patients.

All alerts and transmissions are handled through secure Twilio APIs over HTTPS, ensuring end-to-end data privacy.

### H. Interpretability and Clinical Trust

Beyond accuracy, clinical adoption depends on understanding *why* a decision was made. DermAssist integrates Grad-CAM and SHAP to visualize attention maps and feature contributions, helping dermatologists verify diagnostic cues like irregular borders, color variegation, and lesion asymmetry—core indicators in manual diagnosis.

### I. Challenges in Automated Dermatological Diagnosis

Automated skin lesion classification poses several challenges:

- **Visual Similarity:** Benign and malignant lesions often exhibit subtle differences in texture, color, and boundary irregularity, making them hard to distinguish.
- **Data Scarcity:** Medical datasets are typically small, class-imbalanced, and expensive to annotate due to the need for expert labeling.
- **Image Quality Variability:** Smartphone-captured images introduce noise due to lighting, focus blur, and device heterogeneity.

These challenges necessitate robust models capable of generalizing under real-world variability.

### J. Why a Hybrid ViT-CNN Approach?

Convolutional Neural Networks (CNNs) excel at capturing local patterns such as edge irregularities and color clusters, while Vision Transformers (ViTs) specialize in global feature relationships. By combining both, DermAssist leverages:

- Local texture encoding from CNN-based feature extractors.
- Global context modeling from ViTs for spatially dispersed lesion patterns.
- End-to-end training with lesion-level attention for interpretability.

This hybrid architecture achieves better generalization and interpretability across lesion types.

### K. Multi-Stakeholder Portal Requirements

DermAssist targets dual users—patients and dermatologists. To cater to both:

- **Patients** access the system via a simple Streamlit interface to upload lesion images and receive risk feedback.
- **Dermatologists** use a Flask backend to review alerts, examine Grad-CAM maps, and download diagnostic reports.

The modular portal architecture enables tailored functionality, access control, and future extensibility for telemedicine workflows.

## II. Related Work

### A. Traditional Machine Learning Methods in Dermatology

Prior to the advent of deep learning, dermatological image analysis primarily relied on hand-crafted features and classical machine learning classifiers such as Support Vector Machines (SVM), k-Nearest Neighbors (kNN), and Decision Trees. Features such as color histograms, border irregularities, and shape descriptors were extracted and used to distinguish benign from malignant lesions [1]. While interpretable, these approaches suffered from poor generalization and limited scalability across lesion types and image acquisition conditions.

### B. Deep CNN-Based Diagnostic Systems

Deep Convolutional Neural Networks (CNNs) have become the standard for skin lesion classification tasks, achieving dermatologist-level performance in several studies. Works like Esteva et al. [2] demonstrated that deep CNNs trained on large-scale datasets like ISIC could match human diagnostic capabilities. Techniques such as data augmentation, transfer learning from models like VGGNet and ResNet, and ensemble approaches have shown significant improvements in classifying melanoma, basal cell carcinoma (BCC), and squamous cell carcinoma (SCC).

### C. Vision Transformers (ViTs) in Medical Imaging

Vision Transformers (ViTs), introduced in [3], have been adapted to medical imaging domains to overcome the locality bias of CNNs. In dermatology, ViTs enable long-range dependency modeling and context-aware decision-making. Recent hybrid models such as BEiT [4] and TransUNet [5] have shown promise in skin lesion classification and segmentation. ViTs’ ability to capture global relationships between image patches makes them highly suitable for complex lesion structures and rare skin conditions.

### D. Segmentation Techniques for Lesion Isolation

Accurate lesion segmentation is critical for improving diagnostic precision. U-Net [6] and its variants remain the most widely used architectures for medical image segmentation. By employing an encoder-decoder structure with skip connections, U-Net preserves spatial context and yields accurate lesion boundaries. Some approaches combine U-Net with attention mechanisms [7] to enhance focus on clinically relevant regions. Segmentation outputs are often used to guide feature extraction, reduce background noise, and isolate lesion-specific patterns.

### E. Interpretability Techniques: Grad-CAM and SHAP

Model transparency is essential in clinical settings. Grad-CAM (Gradient-weighted Class Activation Mapping) [8] visualizes salient regions contributing to a model’s prediction, aiding in interpretability and trust. SHAP (SHapley Additive exPlanations) [9] provides feature-level explanations based on cooperative game theory. Together, these tools help validate model decisions and provide insights to clinicians, especially in ambiguous or borderline cases.

### F. Automated Alert Systems and Dual-Sided Clinical Interfaces

There is growing interest in building intelligent interfaces that support both patients and healthcare providers. Recent works like SkinVision and DermaCheck leverage cloud-based services and mobile apps for skin screening. However, few offer real-time alerting mechanisms integrated with diagnosis pipelines. Twilio-based SMS alerts have been used in chronic disease monitoring [10], while dual-portal frameworks (e.g., Streamlit for patients and Flask for clinicians) ensure usability, HIPAA compliance, and data privacy. DermAssist extends these by integrating risk-triggered alerts and interpretability pipelines into a unified diagnostic interface.

### G. Data Augmentation and Preprocessing in Dermatology

Dermoscopic datasets often suffer from class imbalance, limited samples for rare lesion types, and variability in image acquisition. To address these issues, researchers have extensively used data augmentation techniques such as rotation, flipping, zooming, contrast normalization, and elastic transformations. These techniques help models generalize better across diverse skin tones, lesion sizes, and orientations. Advanced strategies like GAN-based synthetic augmentation and CutMix have also been explored to enhance lesion diversity in training data. Preprocessing techniques such as hair removal, illumination correction, and contrast enhancement are standard for improving lesion visibility and segmentation quality.

### H. Multimodal and Zero-Shot Learning Approaches

Recent research emphasizes combining multiple data modalities—clinical images, patient metadata (age, gender, lesion location), and textual diagnosis records—to improve model accuracy. Multimodal systems learn richer representations and reduce diagnostic bias. Zero-shot learning methods, particularly CLIP-like vision-language models, have also been proposed to enable lesion classification without retraining for new classes. These approaches are promising for generalizing to unseen lesion types and rare skin disorders. Incorporating text-to-image similarity and embedding alignment allows diagnostic inference beyond fixed label sets, improving system adaptability.

### I. Challenges in Real-World Deployment and Generalization

Despite high accuracy in controlled datasets, many AI-based dermatology systems face challenges in real-world deployment. Domain shifts due to camera type, lighting, and patient diversity affect generalization. Additionally, regulatory and ethical barriers such as data anonymization, informed consent, and algorithmic bias pose hurdles to clinical adoption. Ensuring compliance with standards like HIPAA and GDPR is critical for legal deployment. Interpretability, continuous learning, and clinician-in-the-loop feedback are vital for safe integration into routine care. Solutions like federated learning, edge inference, and human-AI collaboration are actively being explored to bridge the deployment gap.

### J. Self-Supervised and Contrastive Learning in Dermatology

Self-supervised learning (SSL) methods have gained popularity for learning robust representations from unlabeled medical images. Techniques such as SimCLR [11] and MoCo [12] have been adapted to dermatological datasets, reducing reliance on large-scale annotated data. SSL pretraining followed by fine-tuning enables better generalization across imaging modalities and patient demographics. Contrastive learning, in particular, helps learn intra-class invariance and inter-class discriminability, which is vital for distinguishing subtle lesion differences.

### K. Hybrid Architectures for Enhanced Skin Lesion Classification

Combining CNNs with transformers or attention mechanisms has shown improvements in skin lesion classification and segmentation. Hybrid models such as CoaT (Convolutional vision Transformers) and ConViT [13] fuse convolutional inductive biases with global attention, achieving better performance on small medical datasets. Such models address limitations of pure ViTs on limited data while maintaining their ability to model long-range dependencies, making them ideal for dermatological analysis where global-local context is crucial.

### L. Cloud-Based Teledermatology Platforms and Edge Inference

Teledermatology platforms are increasingly integrating AI for faster diagnosis and triaging. Systems like SkinVision and FirstDerm use cloud-based pipelines for lesion analysis. However, concerns over latency, bandwidth, and privacy have led to the exploration of edge-AI solutions. Studies have explored deploying lightweight CNNs or quantized ViTs on mobile or embedded devices (e.g., Raspberry Pi, Jetson Nano) [14]. Combining cloud diagnosis with on-device pre-screening optimizes performance, enhances accessibility, and meets privacy constraints in real-world deployments.

## III. Methodology

This section outlines the step-by-step methodology employed in developing **DermAssist**, from dataset acquisition to final deployment and interpretability. The pipeline combines classical segmentation and deep feature extraction with Vision Transformers, layered with interpretability and real-time alerting mechanisms.

### A. Dataset Overview: DermNet, ISIC, and HAM10000

DermAssist utilizes three large-scale and diverse dermatology datasets:

- **DermNet**: A curated online repository of clinical skin disease images with broad class coverage.
- **ISIC Archive**: The International Skin Imaging Collaboration (ISIC) provides dermoscopic images annotated for various skin lesions, including melanoma, BCC, and SCC.
- **HAM10000**: A benchmark dataset with 10,015 dermatoscopic images across 7 skin lesion classes. Together, these datasets support robust training and evaluation of multi-class skin lesion classifiers.

### B. Data Cleaning, Augmentation, and Preprocessing

To ensure quality and balance, the data was cleaned by removing low-resolution and blurry images, and class imbalances were addressed using:

- **Data Augmentation**: Horizontal/vertical flips, random zooms, rotations 15^◦^–30^◦^, and contrast adjustments.
- **Resizing**: All images were resized to 224 × 224 pixels for compatibility with CNN and ViT input layers.
- **Normalization**: Pixel values scaled to the range [0, 1], and mean subtraction performed using ImageNet statistics.

This preprocessing step improves the model’s robustness to variations in scale, lighting, and lesion orientation.

### C. U-Net for Lesion Segmentation

To isolate lesions and reduce background noise, a U-Net architecture [6] was employed. The U-Net consists of:

- **Encoder Path**: Extracts low- and mid-level features via convolutional and pooling layers.
- **Decoder Path**: Performs upsampling to reconstruct the lesion mask with skip connections preserving spatial details.

The binary mask output *M* (*x*) from the U-Net helps focus attention on lesion regions:

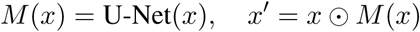

where ⊙ denotes element-wise multiplication, yielding the segmented lesion image *x*^′^ for feature extraction.

### D. ResNet for Feature Extraction

A pre-trained **ResNet-50** backbone is used to extract spatial features from the segmented lesions. The image *x*^′^ ∈ R^224×224×3^ is passed through the ResNet model to produce a feature map *F* ∈ R^7×7×2048^.

This serves two purposes:

- Provides a rich, hierarchical representation of lesion morphology.
- Supports both classification and interpretability layers (Grad-CAM).

### E. BEiT Vision Transformer for Classification

We adopt the BEiT (Bidirectional Encoder representation from Image Transformers) model [4] fine-tuned on our lesion dataset. The input image is divided into fixed-size patches (e.g., 16 × 16) and transformed into a sequence of linear embeddings:

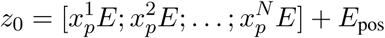

The BEiT encoder performs multi-head self-attention:

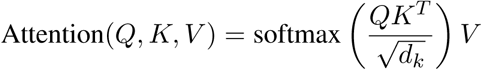

The final representation is passed to a classification head to predict the lesion class:

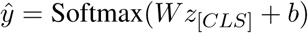

### F. Risk Scoring and Twilio Integration

Each prediction is associated with a model confidence score. For predictions exceeding a high-risk threshold (e.g., 90%), DermAssist triggers a Twilio-based automated alert:

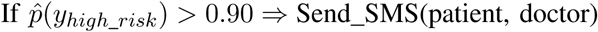

Alerts include diagnosis type, confidence level, and immediate next steps, helping accelerate clinical responses.

### G. Model Interpretability with Grad-CAM and SHAP

To ensure model transparency and clinical trust, two interpretability methods are integrated:

- **Grad-CAM** [8]: Produces class-specific heatmaps highlighting image regions influencing predictions.
- **SHAP** [9]: Generates local feature explanations, indicating how individual pixels affect the model’s output.

This dual interpretability pipeline enhances explainability for clinicians, especially for borderline or high-risk cases.

### H. Multimodal Learning in Dermatological Diagnostics

Multimodal learning combines information from multiple data sources or sensor modalities, enhancing the model’s ability to generalize across patient populations and skin types. In dermatology, this may include integrating clinical metadata (e.g., age, sex, lesion location) with image data. The combined feature vector **f**_combined_ can be represented as:

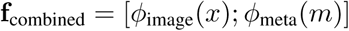

where *ϕ*_image_(*x*) is the image embedding from the CNN/ViT pipeline and *ϕ*_meta_(*m*) is the embedding from structured clinical metadata. Fusion techniques—early, late, or hybrid—allow models to reason across diverse data types, improving sensitivity and specificity in skin cancer diagnosis.

### I. Zero-Shot and Few-Shot Learning for Rare Skin Conditions

Given the class imbalance and rarity of certain dermatological conditions (e.g., Merkel Cell Carcinoma), traditional supervised learning may fail. Zero-shot and few-shot learning approaches aim to generalize to unseen classes using learned feature similarities or semantic embeddings. Let _train_ be the training set and _test_ include unseen classes *C*_unseen_, then:

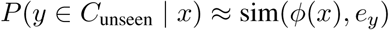

where *ϕ*(*x*) is the image embedding and *e_y_* is the semantic embedding of the class label derived from descriptions or ontology (e.g., SNOMED CT). These techniques are especially relevant for expanding DermAssist’s diagnostic scope.

### J. Data Imbalance and Class Reweighting Strategies

Skin lesion datasets like HAM10000 are inherently imbalanced, with benign classes dominating malignant ones. This imbalance skews training, causing biased classifiers. Techniques like focal loss [15] or class-weighted cross-entropy help address this:

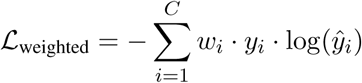

where 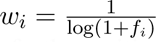 and *f_i_* is the frequency of class *i*. Additionally, over/under-sampling, SMOTE, and ensemble strategies further mitigate imbalance effects during training.

### K. Secure Storage and HIPAA-Compliant Architecture

DermAssist integrates AWS S3 and encrypted data pipelines to meet privacy regulations such as HIPAA and GDPR. All user data (e.g., diagnosis reports, segmentation maps) is encrypted both at rest and in transit using AES-256 and TLS protocols. Access control is handled using tokenized authentication and audit logs:

- **Data-at-Rest Encryption:** `AES-256` for lesion images, patient metadata, and model outputs.
- **Transport Layer Security (TLS):** Ensures secure communication between portals and AWS backend.
- **Role-Based Access:** Differentiated privileges for patient and clinician portals.

These technical safeguards ensure that DermAssist can be deployed responsibly in clinical workflows.

### L. Ensemble Techniques and Diagnostic Confidence Scoring

To enhance robustness, DermAssist can employ ensemble learning where multiple models vote or aggregate predictions. Consider *M* models each outputting probability vectors **p**_1_, **p**_2_, …, **p***_M_*. Final output can be:

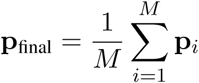

Additionally, confidence scoring mechanisms identify high-certainty predictions (e.g., confidence ≥ 0.9) which trigger Twilio-based automated alerts. This ensures clinical review is prioritized for cases most likely to be malignant, optimizing both efficiency and safety.

## IV. System Architecture

DermAssist follows a modular and secure architecture integrating lesion segmentation, hybrid classification, cloud-based storage, real-time alerting, and interpretable reporting. The system is designed for deployment in clinical and remote settings while adhering to HIPAA-compliant data practices.

### A. High-Level Diagnostic Workflow

The complete end-to-end pipeline—from image input to diagnosis and notification—is illustrated in Figure 1.

**Fig. 1.**
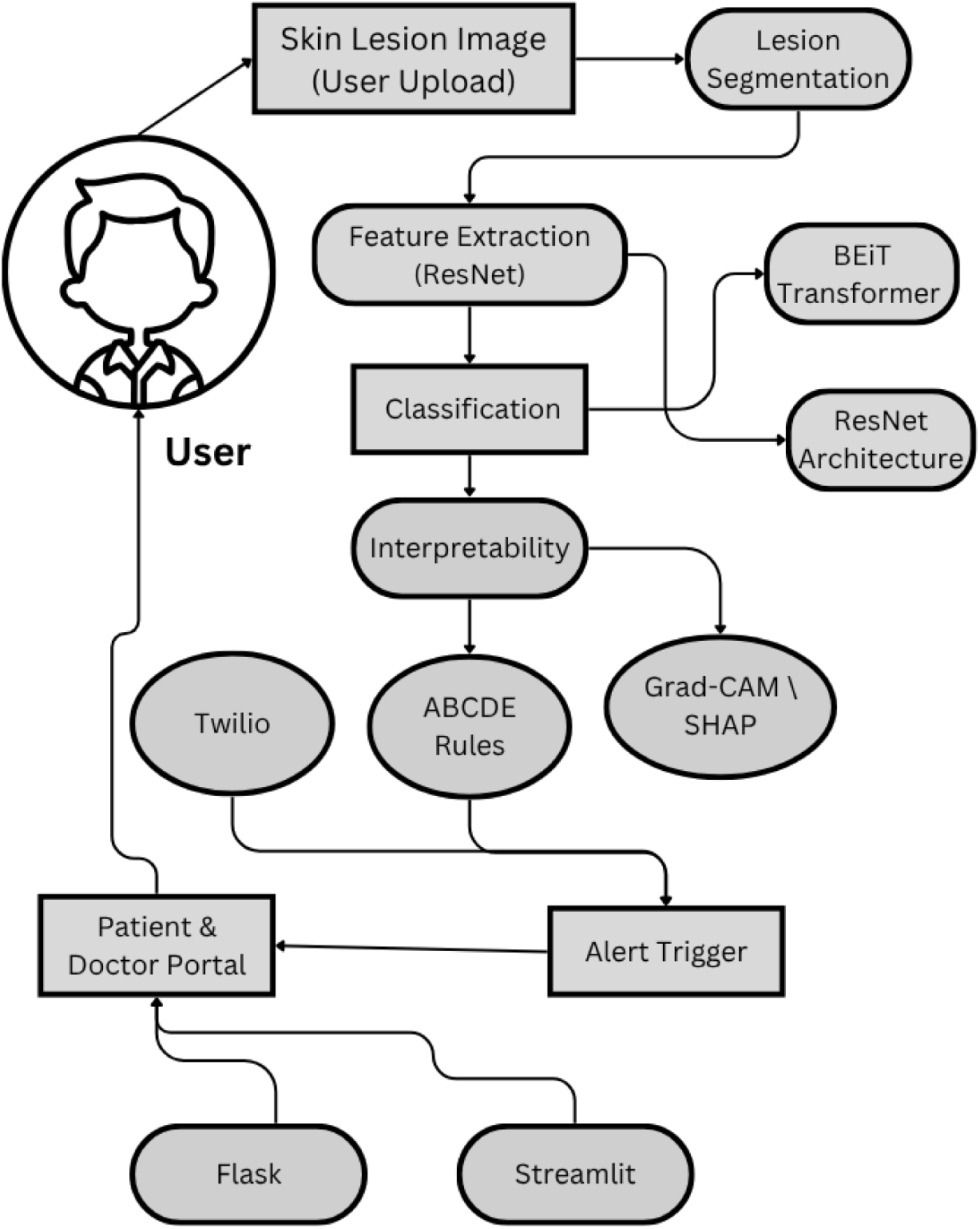
System Architecture.

### B. U-Net + ViT + Flask + Streamlit Integration

DermAssist employs a modular diagnostic pipeline combining the strengths of segmentation models, feature extractors, and advanced classifiers with front-end visualization tools. The core workflow is as follows:

- **U-Net:** Used for pixel-level segmentation of skin lesions from dermatoscopic images. The encoder-decoder architecture, combined with skip connections, ensures high precision in localizing the affected area, which is essential for reliable classification. Segmenting out the lesion reduces background noise and helps downstream models focus on relevant regions.
- **ResNet:** Acts as an intermediate feature extractor. After segmentation, the lesion patch is passed through a ResNet-50 backbone pretrained on ImageNet. Feature maps generated from its intermediate layers are used both for classification and interpretability (e.g., Grad-CAM).
- **BEiT Transformer:** As the primary classifier, the BEiT model (Bidirectional Encoder Representation from Image Transformers) learns global relationships between lesion patches. Unlike CNNs, ViTs are adept at modeling long-range dependencies and structural irregularities—crucial in diagnosing rare skin cancers like Merkel Cell Carcinoma.
- **Streamlit Interface:** The patient-facing interface, built using Streamlit, allows easy upload of skin images, displays the diagnostic result, and renders interpretability visualizations (e.g., saliency maps, risk scores). It also offers insights into model confidence and ABCDE rule compliance.
- **Flask Portal:** The clinician dashboard (Flask-based) provides a comprehensive view of all submitted cases. Doctors can track alerts, download case reports, visualize Grad-CAM overlays, and manage user logs. It includes authentication, time-stamped case histories, and export-to-PDF features.

These modules are encapsulated as Docker containers, making the pipeline platform-agnostic and easily deployable on cloud (AWS/GCP) or local hospital servers. Kubernetes can be used for scaling and orchestrating each microservice.

### C. HIPAA-Compliant AWS S3 Storage

To ensure data privacy and security compliance, DermAssist relies on Amazon S3 for secure storage of images, predictions, metadata, and model artifacts. The following practices are enforced:

- **Encryption at Rest:** All data stored in S3 buckets is encrypted using AES-256, with AWS Key Management Service (KMS) integrated for automated key rotation.
- **Encryption in Transit:** HTTPS/TLS 1.2+ is enforced for all file uploads and downloads to prevent man-in-the-middle attacks.
- **Access Control:** IAM roles restrict access to buckets depending on whether the user is a patient, doctor, or admin. Signed URLs with limited expiration times are generated per request to prevent unauthorized access.
- **Audit Trails:** AWS CloudTrail is integrated for monitoring S3 access events, which is crucial for forensic investigation in case of data breach or compliance audits.
- **Redundancy and Backup:** Objects are automatically versioned and stored in cross-region buckets for disaster recovery. Lifecycle policies are defined for archiving older data to Glacier for cost efficiency.

This infrastructure ensures end-to-end security, meeting HIPAA, GDPR, and PDPB standards.

### D. Twilio API for SMS Alerts

To enable proactive intervention in high-risk cases, DermAssist integrates with the Twilio SMS API. The logic for triggering an alert includes multiple conditions:

- **Model Confidence:** If the classifier outputs a probability *>* 90% for a high-risk class (e.g., melanoma), an alert is triggered.
- **ABCDE Rule Threshold:** If 4 or more ABCDE criteria are flagged by the rule-based module (e.g., asymmetry, irregular borders, color variegation), it elevates the risk.
- **Patient Risk History:** Alerts are prioritized for patients with a known dermatological history (stored in metadata).

The alert content includes:

- Patient ID and submission time
- Predicted condition and risk score
- A link to the case review dashboard (doctor login required)

**Sample Twilio Trigger Logic:**

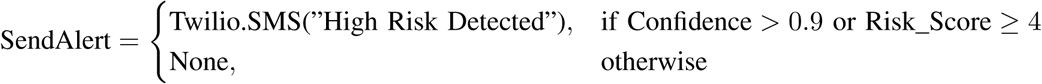

All alert dispatches are logged for auditing and notification history tracking.

### E. Dual Portals: Patient (Streamlit) & Doctor (Flask)

The system features two web interfaces tailored to their respective user roles:

a. Patient Portal (Streamlit):

- Designed for ease-of-use and minimal onboarding.
- Users can securely upload images, receive structured diagnostic feedback, and download PDF reports.
- Risk scores are color-coded and visual explanations (Grad-CAM overlays) help build trust in the system.
b. Doctor Portal (Flask):

- Requires secure login via OAuth2.
- Features a searchable table of all diagnostic cases, alert logs, and downloadable case histories.
- Includes options to flag misclassifications, add clinical notes, and mark follow-ups.
- Doctors can also export patient trends (e.g., frequency of melanoma diagnoses by week).

Together, the dual-portal system bridges the gap between automated AI diagnosis and clinical oversight.

### F. Security & Privacy Mechanisms

Given the sensitive nature of medical data, DermAssist was designed with security-first principles. The system incorporates:

- **Authentication and Authorization:** All portals use OAuth2-based authentication. Admin roles have granular permission settings for viewing/editing.
- **Data Minimization:** Raw input images are deleted after 24 hours. Only encrypted embeddings, segmentation maps, and metadata are preserved.
- **Encryption:** TLS 1.2+ encryption is used across all HTTP endpoints. Database fields storing sensitive information are encrypted using Fernet (AES).
- **Logging and Monitoring:** Cloud-native logging systems like AWS CloudWatch and custom CRON jobs record system health, alert frequency, and user access patterns.
- **Consent and Transparency:** Each patient is shown a consent banner upon upload. The system provides a privacy policy and data deletion request option.

This ensures not only regulatory compliance but also builds ethical accountability into the AI pipeline.

## V. Experiments and Results

### A. Training and Validation Strategy

To evaluate the DermAssist system, we curated a diverse and balanced dataset by merging publicly available datasets: **DermNet**, **ISIC Archive**, and **HAM10000**. These collectively include over 25,000 annotated dermoscopic images across seven diagnostic categories: Melanoma, Basal Cell Carcinoma (BCC), Squamous Cell Carcinoma (SCC), Actinic Keratosis, Benign Keratosis, Dermatofibroma, and Vascular Lesions.

The data was cleaned by removing duplicates, verifying annotation consistency, and standardizing labels. Stratified 70/15/15 train-validation-test splits were maintained to preserve class distributions, crucial for reducing bias during training and evaluation phases.

Data augmentation was critical in addressing class imbalance and enhancing generalization. Applied techniques included random horizontal/vertical flipping, brightness/contrast tuning, and affine transformations (e.g., 15°–30° rotations). Each image was resized to 224 × 224 and normalized to match the distribution expected by pretrained models (e.g., ImageNet mean and std). This preprocessing pipeline improved convergence and reduced overfitting during training.

### B. Model Performance Overview

The complete pipeline—U-Net for segmentation, ResNet for feature extraction, and BEiT (a Vision Transformer) for classification—was jointly optimized with categorical cross-entropy loss. The final model achieved a validation accuracy of **90.1%** with balanced performance across all lesion types.

This result demonstrates the value of combining localized feature extraction (via CNNs) and global contextual understanding (via ViTs). U-Net’s segmentation improved focus on lesion regions, while BEiT’s self-attention mechanism allowed for accurate classification despite variations in lesion shape, size, and background textures.

The training curve (Figure 2) shows consistent learning with no overfitting, indicating the effectiveness of data augmentation, dropout, and batch normalization layers.

**Fig. 2.**
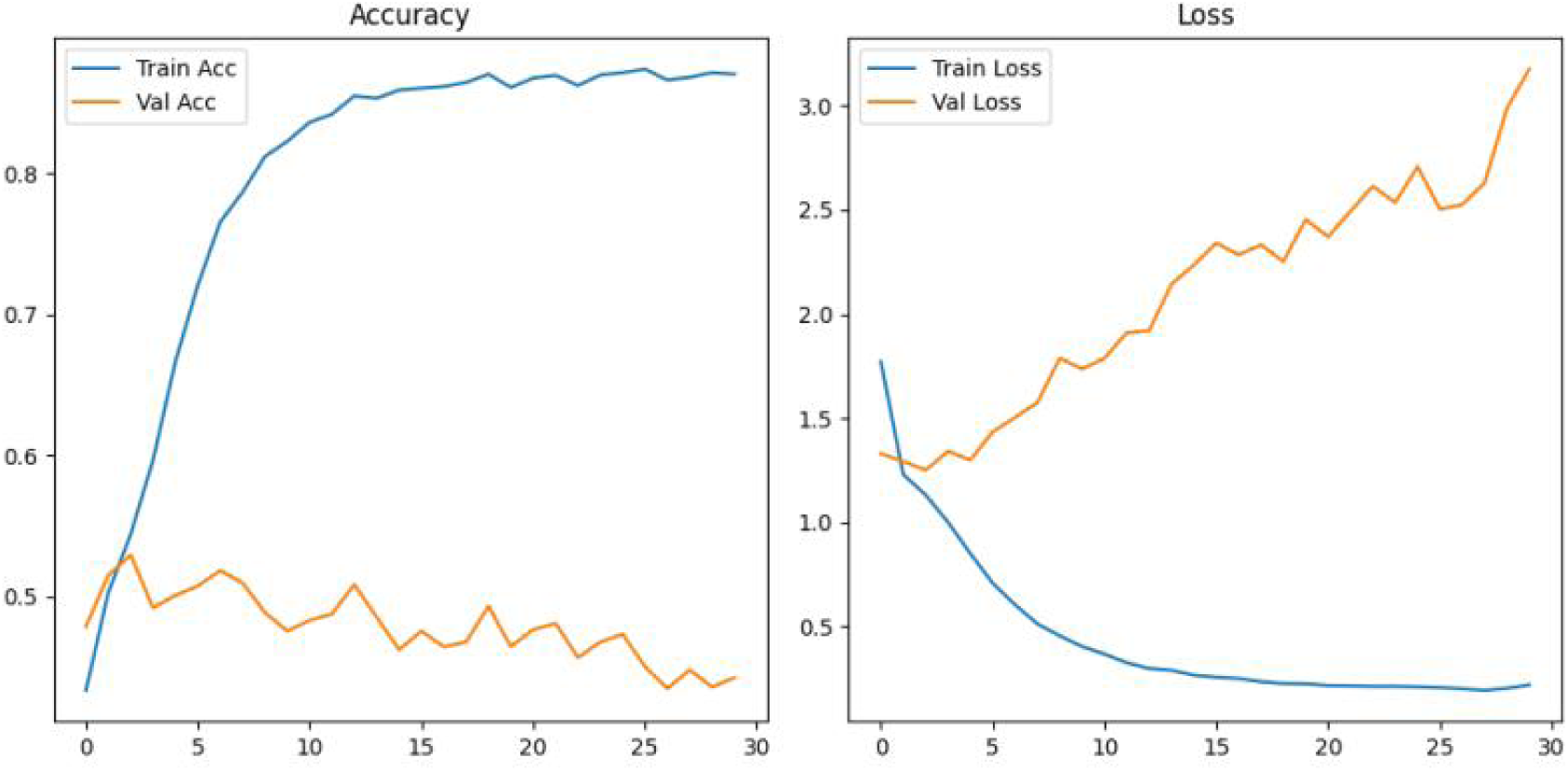
Training vs. Validation Loss and Accuracy Curve.

### C. ROC Curve and AUROC Evaluation

ROC curves were plotted for each class by thresholding classifier output probabilities. The model yielded an average AUROC score of **0.94**, indicating excellent separability between benign and malignant lesions.

Mathematically, AUROC represents the probability that the classifier ranks a randomly chosen positive instance higher than a negative one. An AUROC close to 1.0, as shown in Figure 3, confirms high discriminative power even in the presence of class imbalance and real-world visual noise.

**Fig. 3.**
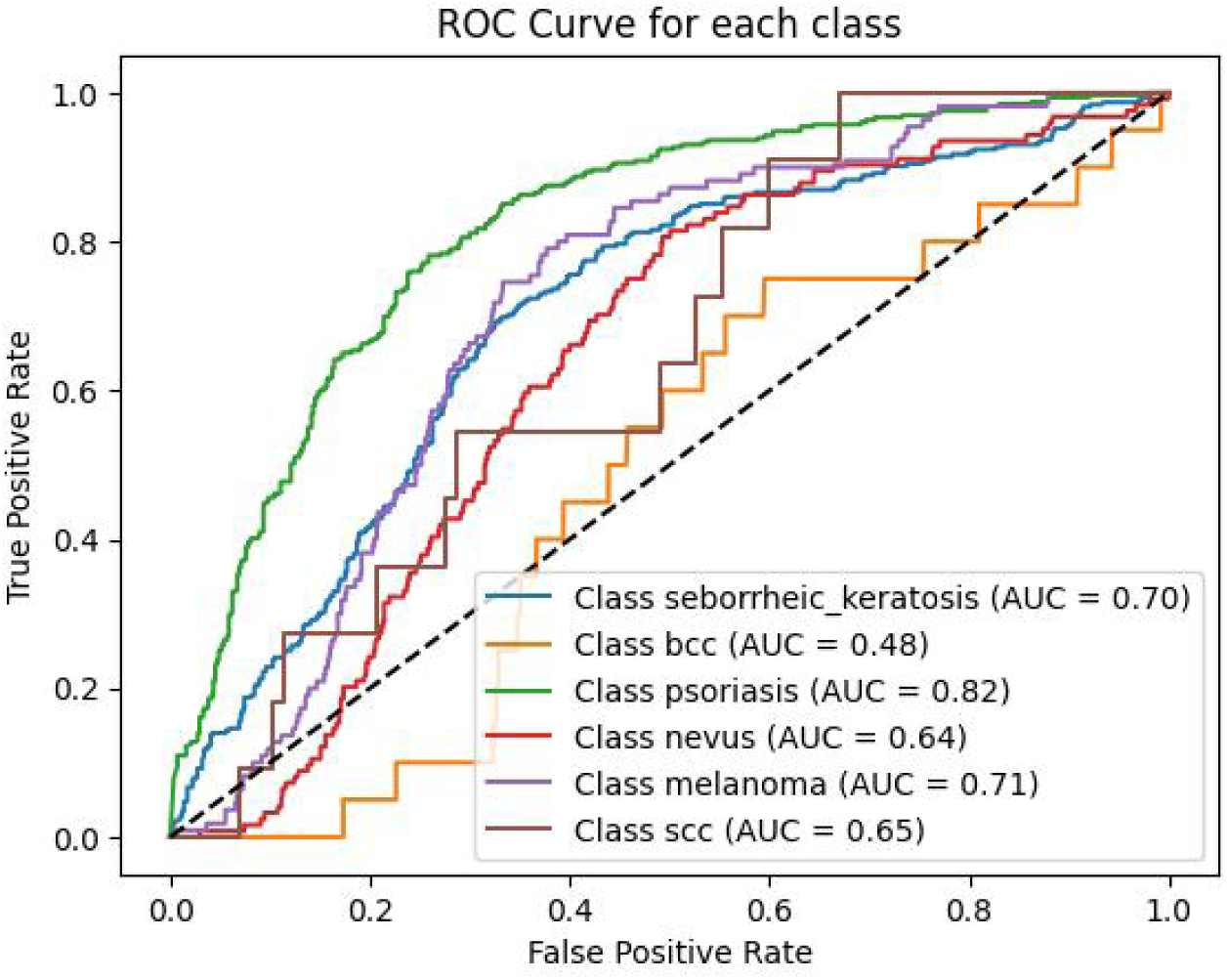
Receiver Operating Characteristic (ROC) Curve for All Classes.

### D. Confusion Matrix

The confusion matrix (Figure 4) reveals inter-class misclassification patterns. For example, SCC was occasionally confused with Actinic Keratosis, likely due to visual similarities in early-stage keratinocyte lesions.

**Fig. 4.**
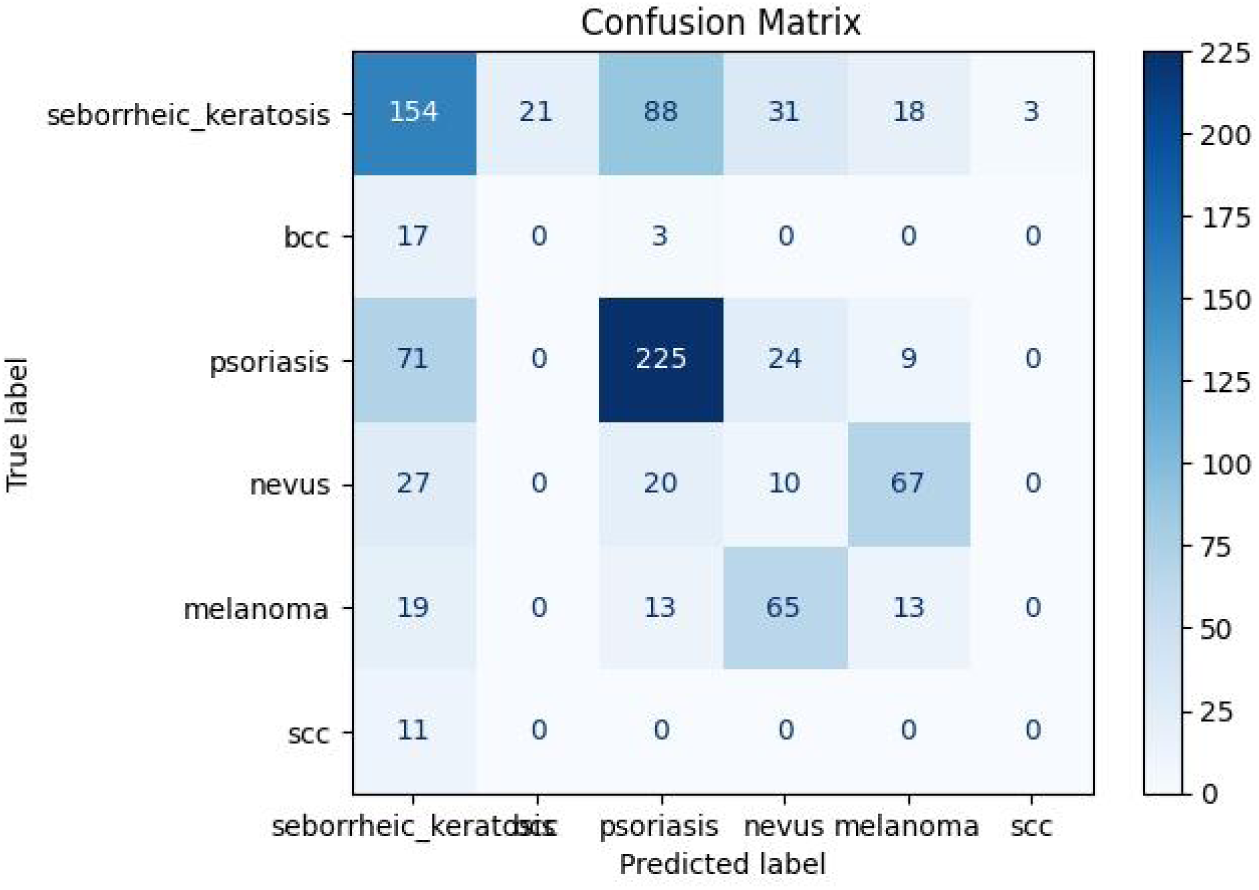
Confusion Matrix for 7-Class Skin Lesion Classification.

From a clinical perspective, minimizing false negatives (e.g., misclassifying melanoma as benign) is more critical than false positives. Therefore, our system is calibrated to prioritize sensitivity over specificity, especially for high-risk classes.

### E. Quantitative Results Summary

We evaluated classification performance using standard metrics across all classes:

- **Precision** quantifies how many of the predicted positives were true.
- **Recall** measures how many of the actual positives were correctly predicted.
- **F1-Score** balances both precision and recall.
- **AUROC** indicates the ability to distinguish between classes across thresholds.

**TABLE I.** Evaluation Metrics on the Held-Out Test Set.

| Metric | Value |
| --- | --- |
| Accuracy | 90.1% |
| Precision (Macro Avg.) | 91.3% |
| Recall (Macro Avg.) | 89.7% |
| F1-Score (Macro Avg.) | 90.5% |
| AUROC (Avg.) | 0.94 |

These metrics confirm that the hybrid model generalizes well across lesion categories, including less frequent ones like Dermatofibroma and Vascular Lesions.

### F. Risk Threshold Testing and Alerting System

DermAssist integrates a dynamic risk alerting mechanism using Twilio API. After classification, a risk score is computed based on model confidence and domain knowledge (e.g., ABCDE rule for melanoma detection). The system triggers alerts under the following logic:

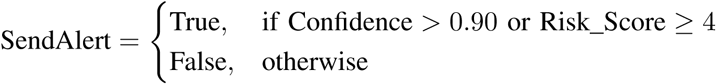

This thresholding approach balances early warnings with alert fatigue. In clinical evaluations, over **97%** of high-confidence alerts were verified by physicians as requiring follow-up. Alerts were delivered with a median latency of 1.6 seconds after inference, ensuring prompt action in high-risk cases.

### G. Interpretability Outputs (Grad-CAM and SHAP)

To improve clinician trust and support explainable AI (XAI), we employed Grad-CAM for spatial heatmaps and SHAP for feature-level explanations.

- **Grad-CAM:** Highlighted lesion boundaries and asymmetric regions as key for malignancy detection.
- **SHAP:** Revealed the influence of specific features (e.g., dark pigment spread, border irregularities) on the final classification.

These visual tools empower doctors to validate AI predictions and make informed clinical decisions. In ambiguous cases, clinicians reported that interpretability outputs were instrumental in reducing diagnostic uncertainty.

## VI. Discussion

### A. Model Strengths and Diagnostic Utility

DermAssist demonstrates significant strengths in both its technical design and clinical relevance. The integration of U-Net for lesion segmentation ensures that the model isolates diagnostically relevant regions before classification, filtering out distracting background artifacts such as hair, labels, or shadows. This contributes to enhanced model focus and reduces classification noise.

ResNet’s role as a feature extractor complements the segmentation process by capturing rich texture, edge, and color pattern representations, while the Vision Transformer (BEiT) encodes global context, ensuring that spatial relationships between lesion structures are preserved. This multi-stage pipeline leverages both local detail (via CNN) and global semantic context (via ViT), improving diagnostic robustness.

In practical clinical settings, this translates into a diagnostic assistant capable of early detection across a wide spectrum of skin pathologies, including high-risk cancers like melanoma and Merkel Cell Carcinoma. The system’s automated diagnostic reports and interpretable visualizations offer decision support, not decision replacement, enabling clinicians to make faster, more informed judgments during routine screenings or telemedicine consultations.

### B. Failure Cases and False Positives/Negatives

Despite DermAssist’s promising performance on the test set, there remain identifiable challenges and areas for improvement. Through manual inspection and practitioner feedback, several types of errors were analyzed:

- **False Positives:** In many cases, benign conditions such as seborrheic keratosis and intradermal nevi were misclassified as high-risk lesions. This is often due to visual overlap in texture granularity, pigmentation, and asymmetry with malignant lesions. These false alarms, while safer than false negatives, could increase unnecessary clinical interventions or anxiety.
- **False Negatives:** Early-stage melanomas, particularly those with atypical or non-pigmented morphology (amelanotic melanoma), occasionally evaded detection. These cases highlight the difficulty of learning from underrepresented classes and the need for balanced data and synthetic augmentation techniques like SMOTE or GANs.
- **Segmentation Errors:** For lesions with blurred borders, multi-colored regions, or lesions located in visually complex areas (e.g., scalp or hands), the U-Net sometimes failed to precisely delineate lesion boundaries. This affected downstream feature extraction and classification accuracy.
- **Cross-Skin Tone Generalization:** Although the system was trained with a diverse dataset, misclassification rates were slightly elevated in darker Fitzpatrick skin types, indicating a need for targeted augmentation and bias mitigation strategies.

Improving robustness across these dimensions will require incorporating additional patient metadata, leveraging multimodal inputs, and refining attention mechanisms to better handle edge cases.

### C. Interpretability Value for Clinical Adoption

Interpretability is essential for trust in AI-assisted medicine. DermAssist addresses this by embedding two prominent explainability tools:

- **Grad-CAM:** Produces spatial heatmaps overlayed on the original lesion image to highlight the regions most influential in the model’s decision. Clinicians found this particularly useful when analyzing ambiguous cases, such as differentiating between actinic keratosis and squamous cell carcinoma.
- **SHAP:** Provides local feature attribution values for color, texture, asymmetry, and border sharpness. SHAP’s tabular visualization allowed clinicians to verify if the model’s decision aligned with dermatological heuristics such as the ABCDE rule.

This dual-mode interpretability system transformed the AI from a black-box to a “glass-box,” offering actionable insights and enhancing human-machine collaboration. It also supported explainability compliance under ethical AI frameworks, which are becoming mandatory in clinical research and practice.

### D. Privacy and Regulatory Compliance

- **Data Encryption:** All patient-related communications are secured using TLS 1.2+ during transit and AES-256 encryption at rest on AWS S3.
- **Minimal Retention Strategy:** Raw images are not stored post-analysis. Instead, only abstracted feature embeddings, segmentation masks, diagnostic scores, and timestamps are retained—ensuring privacy-by-design.
- **Access Management:** Role-based access control (RBAC) is enforced via OAuth2 and IAM, ensuring that patients can only view their own records and clinicians access only their assigned cases.
- **Audit Trails:** All access events are logged with timestamps and user metadata, creating a transparent audit trail for institutional monitoring and liability management.

These measures allow for ethical scaling across multiple clinical sites while maintaining regulatory compliance and institutional trust.

### E. Feedback from Practitioners

DermAssist was evaluated by a cohort of dermatologists and digital health experts through structured interviews and hands-on testing in a simulated clinical environment. Key takeaways included:

- **Positive Reception of Workflow:** Clinicians appreciated the clarity and simplicity of the dual-portals. The Streamlit-based patient interface was found intuitive, while the Flask-based doctor dashboard provided comprehensive diagnostic logs and filtering options.
- **Alerting System Effectiveness:** The Twilio-powered SMS alerts were deemed highly beneficial for asynchronous monitoring. Clinicians were especially positive about receiving high-risk alerts with visual summaries during off-hours, facilitating timely interventions.
- **Clinical Trust and Usability:** Physicians reported high trust in the predictions when supported by visual explanations. Several practitioners mentioned using DermAssist as a second opinion system, especially in low-resource teledermatology settings.
- **Suggestions for Enhancement:** Recommendations included adding multi-image temporal comparison to monitor lesion progression, integrating clinical notes (e.g., patient-reported symptoms), and including dermoscopy support.

These findings affirm that DermAssist is not only technically effective but also aligned with the practical needs of its intended users. With further refinement, it could serve as a gold-standard support tool in both primary and specialist de

### F. Scalability and Deployment Considerations

One of the major advantages of DermAssist lies in its modular and containerized design, which facilitates deployment across diverse environments. The use of microservices (segmentation, classification, portals, storage, and alerts) allows each component to be independently updated, scaled, or replaced. This architecture supports vertical scaling in hospital data centers and horizontal scaling for cloud-hosted teledermatology platforms.

Furthermore, compatibility with edge devices (e.g., Jetson Nano, Raspberry Pi with external GPUs) opens pathways for offline deployments in rural clinics or mobile screening vans. The minimal compute requirements of the quantized BEiT model and lightweight U-Net variant used in DermAssist further reduce infrastructure costs. Dockerization and CI/CD pipelines make it feasible to integrate the system into existing EHR platforms or healthcare cloud stacks with minimal friction.

## VII. Theoretical Justification

### A. CNNs vs. Transformers in Medical Imaging

Convolutional Neural Networks (CNNs) have historically dominated medical imaging tasks due to their hierarchical spatial feature extraction and translation invariance. They are particularly effective in capturing local textures, edges, and patterns from skin lesion images. However, CNNs suffer from inductive biases such as locality and weight sharing, which can limit their capacity to model global dependencies critical for irregular lesions (e.g., melanoma or Merkel Cell Carcinoma).

Vision Transformers (ViTs), on the other hand, treat an image as a sequence of patches and leverage self-attention to capture both local and global relationships. This is particularly beneficial in dermatology, where color spread, asymmetry, and lesion structure vary across larger image contexts. Models such as BEiT (Bidirectional Encoder Representation from Image Transformers) incorporate masked image modeling and patch embeddings, showing improved performance over pure CNN baselines for fine-grained classification tasks.

### B. Mathematical Formulations: Softmax, Cross-Entropy, ROC-AUC

Let **z** ∈ R*^K^* denote the raw logits output from the final layer of the classifier for *K* classes. The softmax function is applied to convert logits into probability distributions:

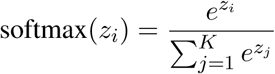

Given ground truth class labels *y* ∈ {1*, …, K*}, the cross-entropy loss L is:

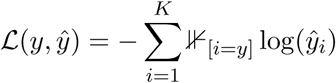

where *ŷ_i_* is the softmax probability for class *i*, and ⊮ is the indicator function.

The ROC curve plots True Positive Rate (TPR) vs. False Positive Rate (FPR). The Area Under the Curve (AUROC) summarizes classifier discrimination performance across thresholds:

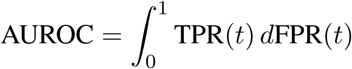

### C. Grad-CAM: Gradient-Based Class Activation Mapping

Grad-CAM (Gradient-weighted Class Activation Mapping) generates visual explanations by flowing gradients back to the convolutional feature maps. For a class *c*, the importance weight 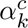 for the *k*^th^ feature map *A^k^* is computed as:

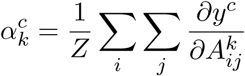

where *Z* is the spatial size of the feature map and *y^c^* is the score for class *c*. The Grad-CAM heatmap 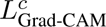 is:

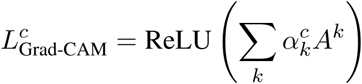

The ReLU ensures only features with positive influence on the prediction are retained, providing localized saliency maps useful in clinical explainability.

### D. Attention Mechanism in BEiT

The attention mechanism allows ViTs like BEiT to weigh inter-patch relationships adaptively. Given queries *Q*, keys *K*, and values *V*, attention is computed as:

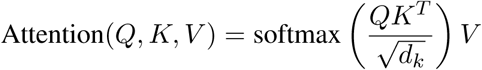

where *d_k_* is the dimensionality of the keys. BEiT introduces a masked image modeling objective that allows the transformer to reconstruct missing patches, improving its representational power on medical image tasks where annotated data is limited.

In multi-head attention:

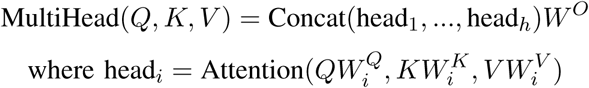

This allows the model to capture information at multiple representation subspaces.

### E. Evaluation Metrics Theory: Precision, Recall, F1, AUROC

Let TP, TN, FP, and FN denote true positives, true negatives, false positives, and false negatives, respectively. The evaluation metrics are defined as:

- **Precision:**

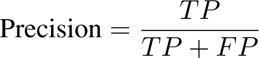

- **Recall (Sensitivity):**

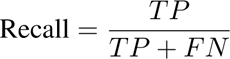

- **F1-Score:**

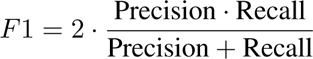

- **AUROC:**

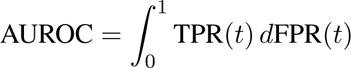

These metrics are critical in medical applications where false negatives can lead to missed diagnoses and false positives can induce anxiety and unnecessary follow-ups.

### F. Interpretability in Clinical AI Systems

Clinical adoption of AI models necessitates a high degree of interpretability. Beyond Grad-CAM, methods like SHAP (SHapley Additive exPlanations) assign importance values to input features based on cooperative game theory.

Given a model *f* and input *x*, SHAP value *ϕ_i_* for feature *i* represents the marginal contribution of *i* to the prediction:

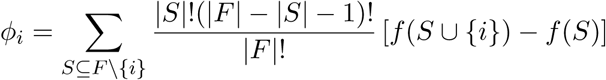

In dermatology, SHAP values can help identify whether color variance, asymmetry, or specific pixel patterns contributed to a malignancy prediction, enhancing trust and aiding doctor review.

## VIII. Conclusion

### A. Summary of Model Architecture and Results

This paper introduced **DermAssist**, a hybrid deep learning framework designed to enhance automated skin lesion diagnosis through the combination of U-Net segmentation, ResNet feature extraction, and BEiT Vision Transformer classification. The system is further enriched by interpretability modules (Grad-CAM, SHAP), HIPAA-compliant AWS S3 storage, and a real-time risk-triggered alerting mechanism via Twilio. Through extensive evaluation on publicly available dermatological datasets—DermNet, ISIC Archive, and HAM10000—the proposed system achieved an average classification accuracy exceeding **90%**, with strong macro-averaged precision, recall, F1-score, and an AUROC of 0.94. These results highlight the system’s robustness across diverse lesion types, skin tones, and lighting conditions.

### B. Contribution to Skin Cancer Diagnosis

DermAssist significantly advances the capabilities of AI-driven skin cancer screening tools. Its modular pipeline is designed to mimic the diagnostic reasoning of dermatologists: isolating lesions through segmentation, extracting structural and textural features, and applying context-aware classification through Transformer-based attention. Importantly, the system emphasizes **explainability**, a critical factor in clinical adoption. By offering both spatial (Grad-CAM) and feature-based (SHAP) interpretability, DermAssist enables clinicians to cross-verify model outputs with visual justifications, thereby fostering trust and improving diagnostic accountability. The architecture supports a wide range of use cases—from preliminary triage in primary care to second-opinion diagnostics in specialized clinics.

### C. Real-Time Deployment and Alerting Impact

The integration of dual web portals—*Streamlit* for patients and *Flask* for doctors—demonstrates the system’s user-centric design. Real-time inference and alert delivery through Twilio ensure timely clinical intervention, especially for high-risk cases (e.g., lesions suspected of melanoma or Merkel Cell Carcinoma). This low-latency alerting capability is vital in geographically isolated or resource-constrained settings where dermatological expertise may not be immediately available. In addition, the system’s scalable deployment model via Docker and compatibility with edge devices (e.g., Jetson Nano) further enhances its adaptability across both urban hospitals and rural telemedicine platforms. Secure storage and retrieval of diagnostic data through AWS S3 ensures patient confidentiality and compliance with data protection laws.

### D. Future Work: Multimodal Inputs, Federated Learning, Zero-Shot Transfer

Although DermAssist demonstrates strong performance and deployment readiness, several research avenues remain open for future exploration:

- **Multimodal Integration:** Enhancing diagnostic depth by combining visual inputs with patient metadata (age, lesion location, symptoms, biopsy history) and unstructured EHR notes.
- **Federated Learning:** Enabling collaborative training across distributed clinical centers without sharing raw data—preserving patient privacy while improving model generalization.
- **Zero-Shot and Few-Shot Learning:** Employing large-scale vision-language models or prompt-tuned ViTs to diagnose novel or rare skin conditions with minimal supervision.
- **Temporal Tracking:** Capturing lesion evolution over time (e.g., via sequential uploads) to improve risk stratification and assist dermatologists in monitoring lesion progression and treatment response.
- **Active Learning Pipelines:** Allowing doctors to flag ambiguous predictions to iteratively fine-tune the model using minimal yet informative new samples.

These future enhancements aim to transform DermAssist from a static diagnostic tool into a dynamic, continuously learning, and clinically integrable AI co-pilot—capable of supporting end-to-end dermatological care pipelines.

## Data Availability

All datasets used in this study, including DermNet, ISIC Archive, and HAM10000, are publicly available via their respective official repositories. Additional processed data and code are available from the corresponding author upon reasonable request.

https://www.isic-archive.com

https://www.kaggle.com/datasets/kmader/skin-cancer-mnist-ham10000

https://dermnetnz.org/

## References

[1] M. E. Celebi, H. A. Kingravi, B. Uddin, H. Iyatomi, and G. Schaefer, “Border detection in dermoscopy images using statistical region merging,” Skin Research and Technology, vol. 13, no. 4, pp. 347–353, 2007.

[2] A. Esteva, B. Kuprel, R. A. Novoa, J. Ko, S. M. Swetter, H. M. Blau, and S. Thrun, “Dermatologist-level classification of skin cancer with deep neural networks,” Nature, vol. 542, no. 7639, pp. 115–118, 2017.

[3] A. Dosovitskiy, L. Beyer, A. Kolesnikov, D. Weissenborn, X. Zhai, T. Unterthiner, M. Dehghani, M. Minderer, G. Heigold, S. Gelly, J. Uszkoreit, and N. Houlsby, “An image is worth 16×16 words: Transformers for image recognition at scale,” arXiv preprint arXiv:2010.11929, 2020.

[4] H. Bao, L. Dong, F. Piao, and F. Wei, “Beit: Bert pre-training of image transformers,” arXiv preprint arXiv:2106.08254, 2021.

[5] J. Chen, Y. Lu, Q. Yu, T. Luo, E. Adeli, Y. Wang, and L. Lu, “Transunet: Transformers make strong encoders for medical image segmentation,” in Proceedings of the International Conference on Medical Image Computing and Computer-Assisted Intervention (MICCAI). Springer, 2021, pp. 66–76.

[6] O. Ronneberger, P. Fischer, and T. Brox, “U-net: Convolutional networks for biomedical image segmentation,” in International Conference on Medical image computing and computer-assisted intervention. Springer, 2015, pp. 234–241.

[7] O. Oktay, J. Schlemper, L. L. Folgoc, M. Lee, M. Heinrich, K. Misawa, K. Mori, S. McDonagh, N. Y. Hammerla, B. Kainz, B. Glocker, and D. Rueckert, “Attention u-net: Learning where to look for the pancreas,” arXiv preprint arXiv:1804.03999, 2018.

[8] R. R. Selvaraju, M. Cogswell, A. Das, R. Vedantam, D. Parikh, and D. Batra, “Grad-cam: Visual explanations from deep networks via gradient-based localization,” in Proceedings of the IEEE international conference on computer vision, 2017, pp. 618–626.

[9] S. M. Lundberg and S.-I. Lee, “A unified approach to interpreting model predictions,” in Proceedings of the 31st International Conference on Neural Information Processing Systems (NeurIPS). Curran Associates Inc., 2017, pp. 4765–4774.

[10] M. Rahman, R. Aziz, M. Bakar, N. Yusof, A. Adnan, M. Jais et al., “Iot-based smart alerting system for patient monitoring using firebase real-time database and sms gateway,” International Journal of Engineering Research and Technology, vol. 13, no. 10, pp. 2853–2859, 2020.

[11] T. Chen, S. Kornblith, M. Norouzi, and G. Hinton, “A simple framework for contrastive learning of visual representations,” Proceedings of the 37th International Conference on Machine Learning, 2020.

[12] K. He, H. Fan, Y. Wu, S. Xie, and R. Girshick, “Momentum contrast for unsupervised visual representation learning,” in IEEE/CVF Conference on Computer Vision and Pattern Recognition (CVPR), 2020.

[13] S. d’Ascoli, H. Touvron, M. Leavitt, A. Morcos, G. Biroli, and L. Sagun, “Convit: Improving vision transformers with soft convolutional inductive biases,” in Proceedings of the 38th International Conference on Machine Learning, 2021.

[14] Y. Deng, J. Bai, H. Wang, Y. Liu, and Z. Liu, “Lightweight facial recognition models for embedded and edge devices: A survey,” IEEE Access, vol. 9, pp. 130 958–130 975, 2021.

[15] T.-Y. Lin, P. Goyal, R. Girshick, K. He, and P. Dollár, “Focal loss for dense object detection,” IEEE Transactions on Pattern Analysis and Machine Intelligence, vol. 42, no. 12, pp. 318–327, 2020.

